# A validation study of the identification of haemophagocytic lymphohistiocytosis in England using population-based health data

**DOI:** 10.1101/2021.05.18.21257169

**Authors:** Mark Bishton, Peter Stilwell, Tim R Card, Peter Lanyon, Lu Ban, Lucy Elliss-Brookes, Jessica Manson, Vasanta Nanduri, Kate Earp, Luke Flower, Raj Amarnani, Judith Rankin, Ethan S Sen, Rachel S Tattersall, Colin J Crooks, Jeanette Aston, Veronika Siskova, Joe West, Mary Bythell

## Abstract

We assessed the validity of coded health care data to identify cases of haemophagocytic lymphohistiocytosis (HLH). Hospital Episode Statistics (HES) identified 127 cases within five hospital Trusts 2013-2018 using ICD-10 codes D76.1, D76.2 and D76.3. Hospital records were reviewed to validate diagnoses. 73/74 patients with confirmed/probable HLH were coded D76.1 or D76.2 (positive predictive value 89.0% [95% CI 80.2-94.9%]). For cases considered not HLH, 44/53 were coded D76.3 (negative predictive value 97.8% [95% CI 88.2%-99.9%]). D76.1 or D76.2 had 68% sensitivity in detecting HLH compared to an established active case finding HLH register in Sheffield. Office for National Statistics (ONS) mortality data (2003-2018) identified 698 patients coded D76.1, D76.2 and D76.3 on death certificates. 541 were coded D76.1 or D76.2 of whom 524(96.9%) had HLH in the free text cause of death. Of 157 coded D76.3, 66(42.0%) had HLH in free text.

D76.1 and D76.2 codes reliably identify HLH cases, and provide a lower bound on incidence. Non-concordance between D76.3 and HLH excludes D76.3 as an ascertainment source from HES. Our results suggest electronic health care data in England can enable population wide registration and analysis of HLH for future research.

## Introduction

Haemophagocytic lymphohistiocytosis (HLH) is a rare syndrome of immune dysregulation characterised by fever, hepatosplenomegaly, cytopenias and the finding of activated macrophages in affected organs (1). Primary HLH is associated with inherited deficiencies of cytotoxic T-cell and NK cell function, while secondary HLH has additional underlying causes include viral and other infections, immunosuppression, auto-immunity or haematological cancers (2, 3). In the context of HLH induced by auto-immunity or inflammation, the term macrophage activation syndrome (MAS) is frequently used as an interchangeable term (4). Mortality rates are high in all patient groups, particularly those with underlying malignancy (5-7) and prompt treatment is critical. HLH is rare with variable clinical features and, apart from primary genetic HLH, lacks validated diagnostic criteria meaning diagnosis relies on clinical pattern recognition and this frequently results in delayed diagnosis and under-estimation of case numbers (8).

Within the UK, as for most countries, it has previously only been possible to carry out regional or tertiary referral-based incidence and survival studies for rare disease such as HLH (6, 7). As HLH may present to several different specialities, such as haematology or rheumatology, previous studies have inherent bias towards specialty-specific diagnostics and consideration of underlying causes potentially resulting in under ascertainment and/or biased conclusions regarding aetiology. Despite the diagnostic and clinical issues that surround HLH, it can be reasonably assumed that all diagnosed cases of HLH would be identified by and treated in secondary care and that diagnostic coding is restricted to a small number of the International Classification of Diseases, 10th Revision (ICD-10) codes (D76.1, D76.2 and D76.3) which should be robust enough to support their use in the identification of cases (9).

The National Congenital Anomaly and Rare Disease Registration Service (NCARDRS) is developing a national register of rare disease for England. The aim of this study was to validate routinely collected hospital admissions and national mortality coding data, against clinically reported data to determine a method to reliably identify patients diagnosed with HLH. Accurate case ascertainment would allow the establishment of a sustainable, ongoing, national, population-based registration of HLH by NCARDRS, resulting in high-quality data adding to the understanding of this rare and often fatal disease.

## Methods

All data were accessed and processed under section 251 of the NHS Service Act 2006. The section 251 permission granted to NCARDRS (CAG 10-02(d)/2015) is the only mechanism by which confidential patient information can be transferred without the discloser being in breach of the common law duty of confidentiality. NCARDRS enabled access to hospital records as well as the national routinely collected healthcare datasets, including Admitted Patient Care (APC) Hospital Episode Statistics (HES) and the Office for National Statistics (ONS) mortality data. HES contains every episode of admitted NHS patient care in England (in-patient and day-case), with all prevalent diagnoses coded ICD-10. ONS mortality data contains both free text and coded cause of death fields.

### Validating HES coding

Potential cases of HLH were extracted from HES using ICD-10 codes D76.1 (Haemophagocytic lymphohistiocytosis), D76.2 (Haemophagocytic syndrome, infection-associated) and D76.3 (Other histiocytosis syndromes) in any diagnostic field for all patients admitted to hospital from 1^st^ January 2013 to 31^st^ December 2018 for five NHS hospital Trusts in England. The trusts were identified through the project team, the UK Hyperinflammation and HLH Across Speciality Collaboration (HiHASC) network and NCARDRS’ remote access permissions to electronic patient records. Data sharing agreements between NCARDRS and these NHS Trusts in England allowed patient details to be made available to expert clinicians in each Trust. The HES extract was further restricted to first admissions with the D76 code data sharing agreements between NCARDRS and these NHS Trusts in England. For four of the hospital Trusts, in-house physicians experienced in the diagnosis and treatment of HLH retrospectively reviewed the clinical, radiological and laboratory results from electronic or physical medical records about these patients and in the fifth, an NCARDRS registration officer with a nursing background reviewed the electronic patient records remotely. Data were extracted via a standardised questionnaire electronic case report form and included information on age at diagnosis, primary vs secondary HLH, co-morbidities, the use of immunosuppressive medications, indices comprising the HLH-2004 diagnostic criteria, the presence or absence of specific infections or underlying cancer and therapies administered including chemotherapy. These parameters, in addition to clinical judgement, were used to decide whether each patient had confirmed, probable, or no evidence of an HLH diagnosis. Confirmed cases of HLH met the HLH 2004 criteria(10), or had a high H-score(11). Cases which didn’t meet either criteria, but had a clear underlying secondary cause associated with HLH and where the expert clinical review felt HLH was the underlying diagnosis were also considered confirmed. Other cases with a strong clinical suspicion who didn’t reach these thresholds were considered probable. In the trust where validation was done by NCARDRS staff, the certainty of diagnosis was determined by accessing the clinic letters. If HLH was given a diagnosis without equivocation, it was considered confirmed. If the letters mentioned HLH as likely or suspected, it was indicated as probable.

For one NHS Trust, a regular HLH multi-disciplinary advisory group (MDAG) meeting had been in place since 2012, with a database recording those patients discussed and diagnosed with HLH. This list was scrutinised by the same physician who validated the HES cases, in order to identify if there were any potential HLH cases not identified using D76 coding and estimate the sensitivity of the HES coding.

### Validating ONS mortality data

Deaths that occurred between 1^st^ January 2003 – 31^st^ December 2018 with ICD-10 codes D76.1, D76.2 and D76.3 occurring anywhere in the coded cause of death fields (parts 1 and 2) were extracted from the ONS mortality data.

The cause of death free text was then searched using text strings (“lymphohist”, “cytic syndrome”, “phago”, “macroph”, “lange”, “cytosis x”, “xantho”, “chester” and “pulmonary hist”) to check for evidence of HLH or MAS or excluding non-HLH conditions with potentially related codes in the free text fields. All records where the text string search returned a result were then checked manually for accuracy. All remaining records were checked manually to determine if they should be classified as HLH, MAS or neither.

### Analysis

Standard summary statistics were used to describe clinical and demographic characteristics of the study cohort. Sensitivity, specificity, PPV, NPV and associated 95% exact binomial confidence intervals were calculated using the “epiR” package (epiR reference) in “R” (version 4.0.5)(12, 13)

## Results

### Hospital Episode Statistics

HES data extraction identified a total of 132 potential cases within the five selected NHS Trusts. These cases were validated against hospital clinical records and a clinical diagnosis of confirmed, probable or not HLH was determined in 127 cases, with clinical and laboratory data obtained where possible. No records were available for the remaining five cases. Following review of these patients, there were a total of 74 cases of confirmed or probable HLH and 53 cases considered not to have HLH (table 1 For those patients with confirmed or probable HLH, 73/74 (98.6%) patients were coded with D76.1 or D76.2 with only one patient with HLH coded with D76.3. Amongst all 82 patients with D76.1 or D76.2 coding 73/82 were confirmed or probable HLH (positive predictive value (PPV) 89.0% [95% CI 80.2-94.9%]). For those patients considered not to have HLH, a total of 44/53 (83.0%) patients were coded with D76.3. Amongst all 45 patients with a D76.3 code 44/45 patients were not considered to have HLH (negative predictive value (NPV) 97.8% [95% CI 88.2%-99.9%]).

**Table 1.**
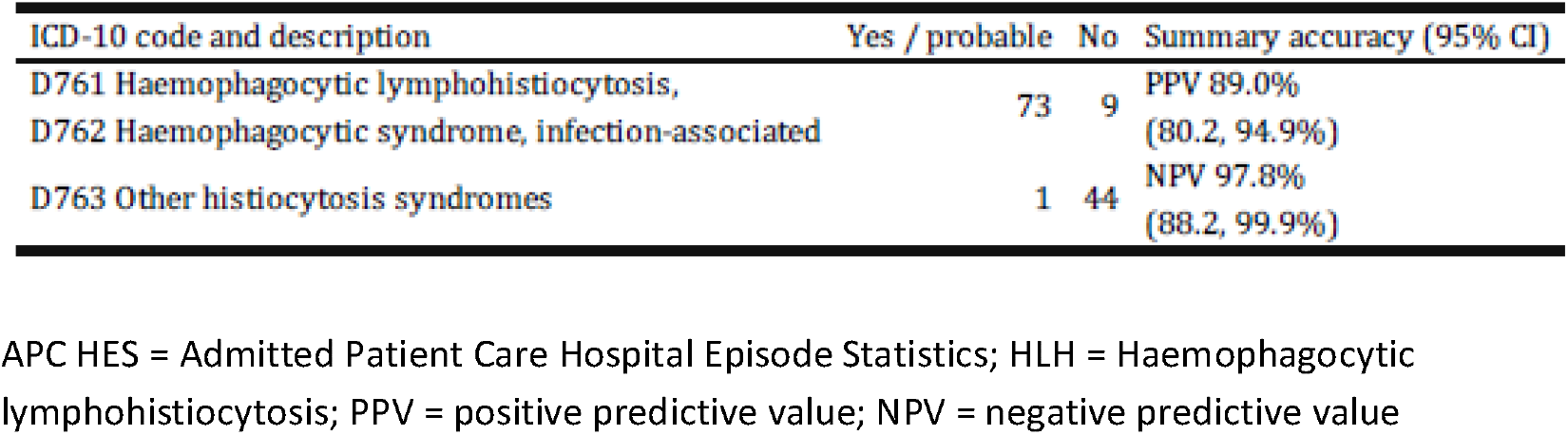
Validation of HES ICD-10 diagnostic coding for HLH in 5 hospital Hemophagocytic lymphohistiocytosis - APC HES data

The patient demographics and laboratory results for those patients with confirmed or probable HLH are summarised in table 2. For the 63/74 (85.1%) cases where data were available, 100% had a ferritin level >500ug/l. Patients with confirmed HLH had a median age of 47 years (range 15.5-59.6) with a male preponderance (61.4%). When factors used to calculate the HLH-2004 score (10) were considered, >90% cases where data were available had documented fevers, cytopenias and elevated triglycerides, and 77.8% had histological evidence of haemophagocytosis. There was no data reported on the presence of fever in 27%, haemophagocytosis in 30% and hepato-or splenomegaly in 33% of confirmed cases. Data collection was better for the laboratory parameters, with only 8-18% missing data for each of the variables. By contrast, patients with probable HLH had a younger median age of 24.7 years (range 16.5-48.6), and fewer males (41.2%). Documented fevers, anaemia and elevated triglycerides were present in >90% cases, and 37.5% of cases had histological evidence of haemophagocytosis. For all the parameters collected, missing data was only present in <10% of probable cases.

**Table 2.** Clinical description of validated cases

| Parameter, % (N), Unavailable % {Unavailable N} * | Confirmed HLH, N = 57 | Probable/Suspected HLH, N = 17 |
| --- | --- | --- |
| Median Age, Years (IQR) | 47 (15.5 - 59.6) | 24.7 (16.5 - 48.6) |
| Male | 61.4 (35) | 41.2 (7) |
| Fever $\geq 38^{\circ}\text{C}$ | 96.7 (29), 47.4 {27} | 92.3 (12), 23.5 {4} |
| Hepato- or splenomegaly | 75 (18), 57.9 {33} | 54.5 (6), 35.3 {6} |
| Hb $\leq 90$ g/l | 97.9 (46), 17.5 {10} | 90.9 (10), 35.3 {6} |
| Platelets $\leq 100 \times 10^9/\text{l}$ | 93.2 (41), 22.8 {13} | 76.9 (10), 23.5 {4} |
| Neutrophils $\leq 1.0 \times 10^9/\text{l}$ | 71.8 (28), 31.6 {18} | 54.5 (6), 35.3 {6} |
| Triglycerides $\geq 3.0$ mMol/l | 92.7 (38), 28.1 {16} | 100 (10), 41.2 {7} |
| Fibrinogen $\leq 1.5$ g/l | 59 (23), 31.6 {18} | 50 (4), 52.9 {9} |
| Ferritin $\geq 500$ $\mu\text{g/l}$ | 100 (49), 14.0 {8} | 100 (14), 17.6 {3} |
| Haemophagocytosis in any tissues | 77.8 (21), 52.6 {30} | 37.5 (3), 52.9 {9} |

\* Unless otherwise stated

### ONS mortality data

ONS mortality data identified a total of 698 deaths in England between 2003-2018 where D76.1, D76.2 and D76.3 appeared anywhere on the death certificates. The ICD-10 cause of death code was compared with the free text validation summary of the death certificate for all patients. Of the 698 patients evaluated, 393 were coded with D76.1, of whom 379 had HLH documented in the free text, 10 had MAS and four had neither. Of the 148 deaths that were coded with D76.2, 145 had HLH documented in the free text, two had MAS and one had neither. Of the 157 patients with D76.3 coding, 66 had HLH documented in the free text, one had MAS, and 90 had neither. Correct classification of HLH/non-HLH increased during the study period from 80.4% [95% CI 75.7%-84.4%] in 2003-2013 to 98.6% [95% CI 96.8%-99.5%] in 2014-2018, after ONS implemented new automated coding software.

Cause of death text strings searches for “lymphohist”, “cytic syndrome”, “phago” or “macroph” correctly classified deaths with HLH or MAS in 599/600 records. Text strings “Lange”, “cytosis x”, “xantho”, “chester” and “pulmonary hist”, correctly excluded 64/64 cases, leaving 34 cases to manually classify. Of the 34 cases manually classified, three were HLH, two MAS and 30 neither.

One of the NHS Trusts involved in the HES validation data maintained a database of patients with HLH proactively identified and recorded since 2012 through an internal HLH Multi-Disciplinary Advisory Group (MDAG). To quantify how many cases of HLH were potentially missed by both HES and ONS D76 mortality coding, the patients on this database were compared with the compiled HES/ONS lists for that Trust. A total of 51 cases of HLH had been recorded, of which 39% (n=20) occurred outside of the study period and 2% (n=1) lacked sufficient information to determine timing. Of the remaining 31 cases, 2168% (n=21) were identified through HES estimating a sensitivity of 67.7% [95% CI 48.6-83.3%], whilst 32% (n=10) of HLH cases were not identified using either HES or mortality data. All HES ICD-10 codes during the calendar year for these 10 cases were identified and examined. The coding most likely to have been used for these episodes were sepsis in four patients (one with an underlying haematological malignancy), and one case each of combined CMV and pneumocystis following chemotherapy, combined TB, CMV and pneumocystis PCP in a patient with an underlying haematological malignancy, juvenile arthritis, perforated duodenal ulcer, systemic lupus erythematosus, and one patient where the underlying diagnosis was unclear.

## Discussion

HLH is a rare and frequently fatal syndrome that can occur at any age, has multiple underlying causes and may present to several different specialities (2). Diseases such as HLH are traditionally difficult to study, but whole population-based studies, using routinely-collected electronic healthcare data via HES and ONS, give an opportunity to identify and validate the ascertainment of electronic healthcare recording of HLH in England.

Overall, we found good ascertainment and PPVs for both HES and death certificates for HLH identification purposes for D76.1 and D76.2. Validation work on HES coding carried out in five separate Trusts using clinical and diagnostic data demonstrated those cases coded with either D76.1 or D76.2 in HES had a high predictive value for HLH. In contrast, the degree of non-concordance between D76.3 and HLH, where only 1/45 cases coded with D76.3 were considered to have HLH, justifies excluding it as a case ascertainment source from HES. Concordance for D76.3 and HLH in the mortality data were lower, but still accounted for a substantial number of deaths attributed to HLH. Concordance improved after 2014 following the implementation of the IRIS, an automated cause of death coding system to code cause of death information provided on death certificates and improve the comparability of mortality statistics (10).

Using a coding approach to identify cases of HLH does result in some limitations. It is unknown how many cases of clinically diagnosed HLH occur which are not coded with any of the D76s, and 10/31 independently annotated HLH cases from one NHS Trust within the study timeframe were not identified by coding. This MDAG has been running for 10 years, is convened between a small group of interested physicians proactively looking for cases of HLH, and likely represents the maximum possible ascertainment process via clinical diagnosis. In addition, identifying cases using only in-patient data could theoretically miss out-patient diagnoses, although the clinical characteristics of HLH means out-patient diagnoses are highly unlikely to be a significant proportion of cases. Our method therefore produces a cohort of patients highly likely to have HLH but we accept likely there will always be consistent under-ascertainment of all possible cases due to the combination of the clinical iceberg effect, variable clinical diagnostic patterns and incorrect coding. Our work provides a standardised method of measuring the incidence and outcomes for this rare disease that currently does not exist and allows the recording of trends in disease, outcomes and response to treatment.

Our methods are likely to improve over time with better clinical ascertainment of cases and clearer use of ICD codes, leading to potential for improvements in care as we determine geographical, social, age, sex and other demographic inequalities in patients.

Other studies have also attempted using coding as a tool to accurately identify cases of HLH. A French study in three tertiary care centres over a period of six years identified all adult patients with any D76 coding, cross-referencing cases with request forms for bone marrow aspirations investigating for HLH as well as lists of all bone marrow aspirations where haemophagocytosis was reported. The clinical and laboratory data for a total of 312 patients was then reviewed by expert investigators and cases were divided into positive (52%), undetermined (14.7%) and negative (33.3%) cases. Notably the proportion of patients in each category was similar to our study; confirmed (44.9%), probable (13.4%) and not HLH (41.7%), as was the median age (47 vs 48 years) and the male preponderance in confirmed cases and female for probable cases, although there was no analysis of the sensitivity or specificity of each D76 code for HLH as described here. A single centre study in the USA also used ICD-9 and ICD-10 codes to identify cases of adult HLH over a 10-year period on the outcomes for 41/50 cases who also met the diagnostic criteria described by Daver et al (5). Of the nine patients excluded from analysis, a total of four had incorrect ICD codes, two were <18 years old at the time of diagnosis, two had been diagnosed at an outside institution, and two had a diagnosis of adult Still’s disease. The findings in these studies appear broadly similar to our own and provide confidence in the use of coding as a means of identification of HLH.

In summary, our results show that using electronic health care data in England is sufficiently reliable to identify cases of HLH and give a reasonably accurate, though underestimated, view of the occurrence of this disease. It also demonstrates the value of collaborative working with NCARDRS under their information governance arrangements. Continued registration of HLH by NCARDRS will allow for the monitoring of long-term incidence trends and patient outcomes, leading to improved understanding of survival, aetiology and treatment options for this rare and often fatal disease.

## Data Availability

All data were accessed and processed under section 251 of the NHS Service Act 2006. The section 251 permission granted to NCARDRS (CAG 10-02(d)/2015) is the only mechanism by which confidential patient information can be transferred without the discloser being in breach of the common law duty of confidentiality in England. patient level data is therefore not available.

## Acknowledgements

Produced by Public Health England. Source for Hospital Episode Statistics (HES) is NHS Digital. Copyright © 2021, Re-used with the permission of the NHS Digital. All rights reserved.

The authors thank Histio UK for funding the project.

